# Detection of Left Ventricular Outflow Obstruction from Standard B-Mode Echocardiogram Videos using Deep Learning

**DOI:** 10.1101/2025.03.02.25323199

**Authors:** Victoria Yuan, Hirotaka Ieki, Christina Binder, Yuki Sahashi, Paul C. Cheng, David Ouyang

## Abstract

**Introduction:** Hypertrophic cardiomyopathy (HCM) affects 20 million individuals globally, with increased risk of sudden death and heart failure. While cardiac myosin inhibitors show great promise as disease-specific treatment, current indications are for obstructive HCM. Obstruction is not always well characterized by echocardiography, and artificial intelligence (AI) might assist in the improving the underdiagnosis of left ventricular outflow tract (LVOT) obstruction.

**Methods:** We identified 2,693 patients with LVOT obstruction and 6,177 control patients matched by age, sex, and septal thickness. A deep learning model was trained on non-Doppler apical-4-chamber (A4C) B-mode echocardiographic videos to detect the presence of outflow obstruction identified later in the same study by spectral Doppler. Model performance was evaluated on held-out test sets from Cedars-Sinai Medical Center (CSMC) and from Stanford Healthcare (SHC).

**Results:** In a held-out test set of 6,034 videos from CSMC, our model demonstrated strong performance in detecting LVOT obstruction with an AUC of 0.865, sensitivity of 77.8%, and specificity of 79.8%. Performance was consistent across various patient subgroups, including those with hyperdynamic LV function, pre-existing valvular disease, and small LV cavity size. The model demonstrated generalizable performance in the SHC cohort with an AUC of 0.866.

**Conclusion:** In this study, we developed an AI model to detect LVOT obstruction from standard A4C echo videos, highlighting patients may benefit from more detailed cardiac workup for obstructive HCM.

## Introduction

Hypertrophic cardiomyopathy (HCM) is an autosomal dominant disease that affects 20 million individuals globally^1^ and increases risk for arrhythmia, sudden cardiac death, and severe heart failure^1–3^. The underlying pathophysiology of obstructive HCM involves myocyte disorganization, systolic anterior motion (SAM) of the mitral valve, and subsequent left ventricular outflow tract (LVOT) obstruction ^4^. Mavacamten, a cardiac myosin inhibitor, was shown in multiple clinical trials to reduce LVOT obstruction, improve heart failure symptoms, and prevent negative cardiac remodelling^5,3,6^. However, LVOT obstruction is frequently missed, with an underdiagnosis rate of nearly 90%^7^. In the United States alone, nearly 600,000 patients are estimated as undiagnosed^4,8^. Given the prevalence of obstructive HCM and its clinical burden, there is a persistently unmet need for earlier disease detection.

Transthoracic echocardiography is the primary tool to diagnose LVOT obstruction; however, evaluation requires careful expert sonographer evaluation and additional images and Doppler measurements for a thorough evaluation. Because LVOT gradients are dynamic, they may also require provocative measures such as Valsalva maneuvers or dobutamine administration to accentuate the gradient for detection. An elevated LVOT gradient is present at rest in only 20% of patients with obstructive HCM, while the rest have LVOT obstruction that becomes apparent only with provocation^9^. Consequently, nearly half of patients with obstructive HCM are missed on resting echocardiography^10,11^. Thus, identifying individuals in standard images who require more detailed assessment can improve detection of obstructive HCM.

Deep learning analysis of echocardiography has the ability to accurately assess cardiac function and detect cardiovascular diseases with high precision^12–17^. AI models have been shown to improve measurement of common parameters such as left ventricular ejection fraction^15^, as well as evaluate echocardiograms for severity of valvular disease^14,16,17^, amyloidosis^12^, and cardiac hypertrophy^12^. Given the challenges in detecting obstructive HCM and the resulting underdiagnosis, machine learning can facilitate early identification of LVOT obstruction. In this study, we developed a deep-learning model that predicts the presence of LVOT obstruction from standard apical-four-chamber (A4C) view videos and evaluated the model in two geographically distinct cohorts.

## Methods

### Study Cohort

Transthoracic echocardiogram studies at Cedars-Sinai Medical Center (CSMC) were evaluated for presence of reported LVOT obstruction. A total of 4,244 studies from 2,693 unique patients who received care between 21 April 2014 and 05 June 2022 were identified to have an LVOT obstruction (Supplementary Figure 1). Patients with systolic motion of the anterior mitral valve without a measured LVOT gradient, patients with an intra- or mid-cavitary gradient, and patients with a gradient induced on provocative testing were also included as case patients. Control patients had no reported LVOT obstruction or gradient and were matched by IVSd in a 2:1 ratio to case patients. Patients with a history of transcatheter aortic valve replacement, post-myomectomy, and studies who did not have measurements for diastolic interventricular septum thickness (IVSd) were excluded from the cohort.

A geographically distinct echocardiography dataset from Stanford Healthcare was identified for external validation of the AI algorithm. Cases consisted of 125 studies from 125 patients with identified LVOT obstruction greater than 50cm/s. 242 studies matched by IVSd from 242 patients matched by age and sex were used as controls. This study was approved by the Institutional Review Boards at Cedars-Sinai Medical Center and Stanford Healthcare.

### Development and evaluation of deep learning model

To construct the deep learning model, the dataset was split using 80% of the dataset for training, 10% for validation, and 10% as a held-out test set to evaluate model performance. Patients were randomized to each set such that there was no crossover between training, validation, and test sets. We trained a binary classifier using an 18-layer R(2+1)D network with a learning rate of 0.01 and Adam optimizer for 26 epochs with early stopping based on validation loss. Model performance was evaluated using confusion matrices, sensitivity, specificity, and area under the receiver operating curve (AUC). Subgroup analysis was conducted to characterize model performance in patients with diverse presentations of obstruction and similar phenotypes. We evaluated the performance of our model on subgroups of patients with mitral regurgitation, aortic stenosis, hyperdynamic LV function and small LV cavity size. Patients with small LV cavity size were identified by analyzing study reports and using study sex-specific thresholds for left ventricular end-diastolic volume index (LVEDV_i_). Similarly, patients with LV hypertrophy were identified through echocardiography reports and sex-specific threshold for septal thickness. We defined patients with hyperdynamic LV as having LVEF ≥ 60%. Patient demographics and comorbidities were extracted from electronic health records using ICD-10 codes. Echocardiography parameters were obtained from study reports.

## Results

From CSMC, 2,693 patients were identified as reported to have LVOT obstruction; the case cohort thus consisted of 4,244 echocardiography studies with 24,931 A4C videos. The control cohort included 6,177 patients matched by IVSd (Kolmogorov-Smirnov test statistic = 0.01, p-value = 0.65) without reported LVOT obstruction, resulting in 8,598 echocardiography studies with 38,094 A4C videos (Supplementary Figure 1). Of the 56% of cases with LVOT gradient measured, 44% had resting gradient < 30 mmHg. Case and control patients had similar age, racial backgrounds, and left atrial volume index (LAVI). Case patients had lower rates of comorbidities such as diabetes, hypertension, atrial fibrillation, and coronary artery disease (Table 1). The data was randomly split on the patient level into training, validation, and test sets (Table 2).

**Figure 1.**
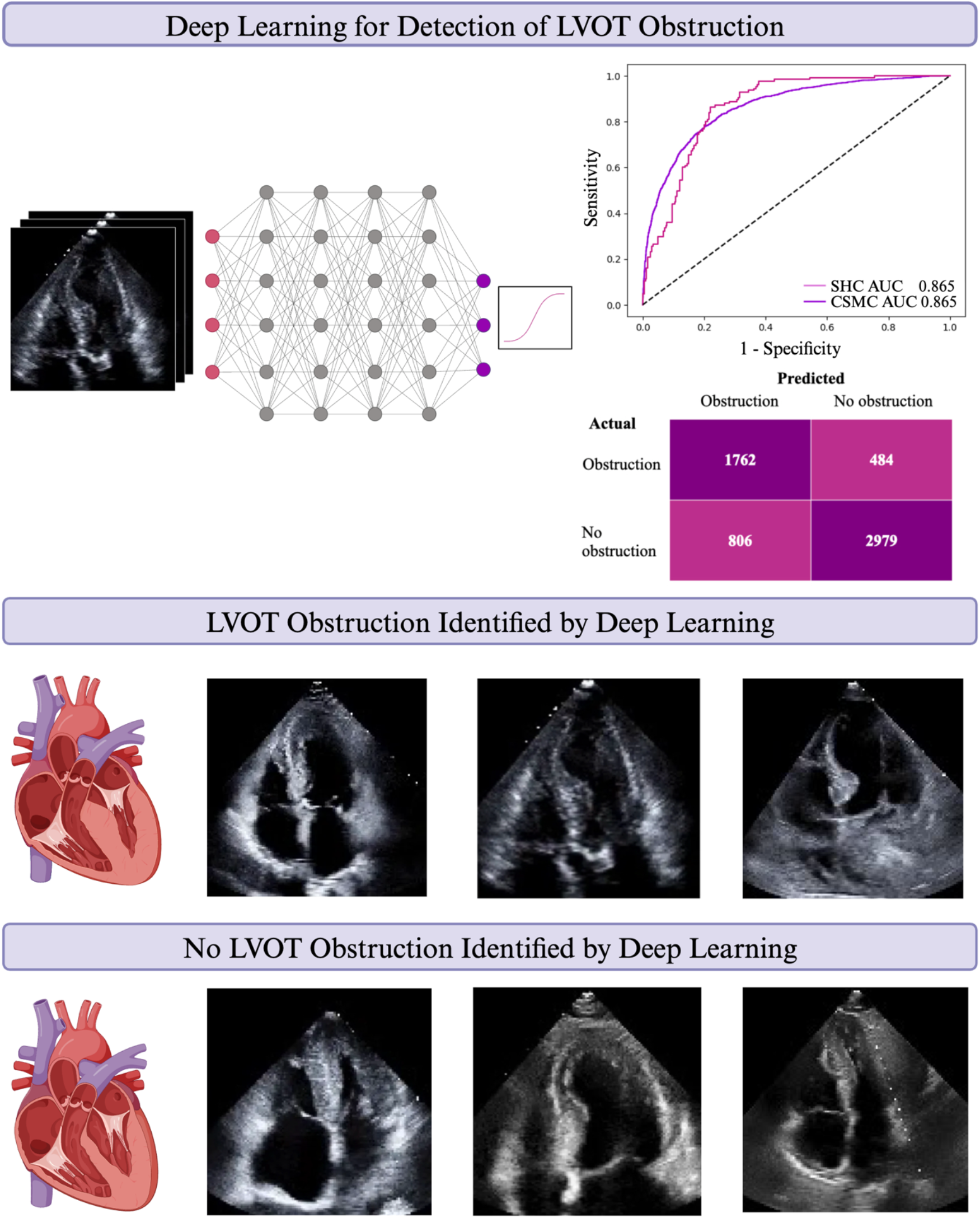
Deep learning algorithm to detect LVOT obstruction from A4C with AUC and confusion matrix (top) Representative apical 4-chamber images for selected DL-predicted cases (middle) and controls (bottom). Created in https://BioRender.com

**Table 1.** Demographic information and comorbidities of study cohort for Cedars-Sinai Medical Center (CSMC) and Stanford Healthcare (SHC)

|  | **CSMC Controls** | **CSMC Cases** | **SHC Controls** | **SHC Cases** |
| --- | --- | --- | --- | --- |
| Age | 69.8±18.0 | 71.5±19.4 | 62.82±17.63 | 62.15±14.16 |
| Sex |  |  |  |  |
| Male | 4,678 (62.4%) | 525 (41.3%) | 160 (66.1%) | 59 (47.2%) |
| Female | 2,782 (37.1%) | 735 (58.1%) | 82 (33.9%) | 66 (52.8%) |
| Race |  |  |  |  |
| American Indian or Alaska Native | 20 (0.27%) | 3 (0.24%) | 0 (0.0%) | 0 (0.0%) |
| Asian | 470 (6.3%) | 93 (7.4%) | 31 (12.8%) | 25 (20.0%) |
| Black or African American | 1,248 (16.6%) | 181 (14.3%) | 30 (12.4%) | 6 (4.8%) |
| Native Hawaiian/Pacific Islander | 25 (0.3%) | 6 (0.47%) | 3 (1.2%) | 0 (0.0%) |
| White | 5,121 (68.29%) | 859 (67.8%) | 116 (47.9%) | 65 (52.0%) |
| Other | 448 (6.0%) | 77 (6.1%) | 41 (16.9%) | 18 (14.4%) |
| Unknown | 167 (2.2%) | 47 (3.7%) | 14 (5.8%) | 11 (8.8%) |
| Ejection fraction,% | 57.4±14.3 | 70.5±24.0 | 55.66±13.56 | 67.37±5.97 |
| Left ventricular end-diastolic volume index (mL/m<sup>2</sup>) | 21.6±18.4 | 11.6±7.3 | 46.68±21.55 | 38.72±11.94 |
| Left ventricular end-systolic volume index (mL/m<sup>2</sup>) | 46.4±24.1 | 36.3±18.2 | 22.44±14.97 | 13.77±4.73 |
| Left atrial dilation | 3,668 (42.3%) | 2,141 (50.6%) | 174 (71.9%) | 104 (83.2%) |
| Diastolic interventricular septal thickness (mm) | 1.39±0.57 | 1.33±0.83 | 1.74±0.41 | 1.78±0.46 |
| Implanted cardioverter-defibrillator | 461 (6.1%) | 19 (0.7%) | 50 (20.7%) | 27 (21.6%) |
| Mitral regurgitation | 545 (7.2%) | 57 (2.1%) | 145 (59.9%) | 98 (78.4%) |
| Coronary artery disease | 2,146 (28.4%) | 183 (6.8%) | 81 (33.5%) | 36 (28.8%) |
| Atrial fibrillation | 2,129 (28.2%) | 207 (7.7%) | 120 (49.6%) | 47 (37.6%) |
| Hypertension | 1,633 (21.6%) | 240 (8.9%) | 149 (61.6%) | 59 (47.2%) |
| Diabetes mellitus | 1,955 (25.9%) | 216 (8.0%) | 97 (40.1%) | 23 (18.4%) |

**Table 2.** Demographic information and comorbidities of train, validation, and test sets

|  | **Train** | **Validation** | **Test** |
| --- | --- | --- | --- |
| Cases | 20,451 (40.1%) | 2,213 (37.2%) | 2,267 (37.2%) |
| Controls | 30,522 (59.9%) | 3,737 (62.8%) | 3,835 (62.8%) |
| Age | 69.8±17.0 | 69.6±16.5 | 71.4±17.3 |
| Sex |  |  |  |
| Male | 26,937 (56.5%) | 3,233 (57.6%) | 2,363 (57.8%) |
| Female | 20,475 (43.0%) | 2,331 (41.5%) | 2,363 (41.1%) |
| Race |  |  |  |
| American Indian or Alaska Native | 89 (0.2%) | 2 (0.03%) | 27 (0.4%) |
| Asian | 3,075 (6.5%) | 410 (6.9%) | 371 (6.2%) |
| Black or African American | 7,354 (15.4%) | 898 (15.2%) | 900 (14.9%) |
| Native Hawaiian/Pacific Islander | 221 (4.6%) | 5 (0.8%) | 16 (2.7%) |
| White | 32,743 (68.7%) | 3,768 (63.8%) | 3,917 (65.9%) |
| Other | 3,298 (6.9%) | 356 (6.0%) | 268 (4.4%) |
| Unknown | 866 (1.8%) | 173 (2.9%) | 121 (2.0%) |
| Ejection fraction (%) | 62.0%±14.6% | 60.83%±14.8% | 62.0±13.7% |
| Left ventricular end-diastolic volume index (mL/m<sup>2</sup>) | 18.6±16.8 | 18.8±15.9 | 17.9±14.2 |
| Left ventricular end-systolic volume index (mL/m<sup>2</sup>) | 43.5±22.0 | 43.3±28.1 | 42.6±18.9 |
| Left atrial volume index (mL/m<sup>2</sup>) | 39.4±22.7 | 39.0±28.7 | 39.3±24.2 |
| Diastolic interventricular septal thickness (mm) | 1.46±0.52 | 1.44±0.43 | 1.47±0.44 |
| Implanted cardioverter-defibrillator | 3,147 (6.2%) | 418 (7.1%) | 358 (5.9%) |
| Mitral regurgitation | 7,849 (15.5%) | 751 (12.7%) | 519 (8.6%) |
| Coronary artery disease | 14,122 (28.0%) | 1,595 (27.0%) | 1,797 (29.8%) |
| Atrial fibrillation | 13,887 (27.5%) | 1,734 (29.4%) | 1,800 (29.8%) |
| Hypertension | 11,765 (23.3%) | 1,485 (25.2%) | 1,478 (24.5%) |
| Diabetes mellitus | 12,448 (24.7%) | 1,456 (24.7%) | 1,518 (25.2%) |

Our model demonstrated strong performance in detecting of LVOT obstruction from standard A4C videos from the same study. In the internal test set, the model showed an AUC 0.865 with a sensitivity of 77.8% and specificity of 79.8% (Figure 1). We also evaluated the model on an external dataset from Stanford Healthcare and the model performed similarly well with an AUC of 0.866, sensitivity of 89.6%, and specificity of 69.4% (Figure 1).

Analyzing patient subgroups further supported model performance. LVOT obstruction was detected across different ranges of LVEF as well as in patients with left ventricular hypertrophy, aortic stenosis, and small LV cavity (Supplementary Figure 2, Supplementary Table 1). We subsequently manually reviewed cases of false positives and false negatives. Of the videos predicted as false-negative – those with physician-reported obstruction but classified by the model as having no obstruction – 30% comprised of patients with inducible obstruction or those with SAM but no obstruction at rest (Supplementary Table 2). Of the false positive videos – those without obstruction and classified by the model as having obstruction – 47% of videos had left ventricular hypertrophy; suggesting further evaluation for LVOT obstruction may beneficial (Supplementary Table 2).

## Discussion

In this work, we developed and validated a deep learning-based model to identify LVOT obstruction from A4C videos, potentially serving as an aid for sonographers to identify patients that should undergo further evaluation for HCM. Our model demonstrated strong performance across diverse patient subgroups and across a range of LVEF, valvular disorders, and different cardiac hypertrophies.

Epidemiological studies emphasize that obstructive HCM is an underrecognized, rather than a rare disease. Patients often present with nonspecific symptoms and interrogation for LVOT obstruction requires additional time and effort during echocardiography. Our findings suggest that deep learning can facilitate efficient recognition of patients who may benefit from further workup. When combined with clinical histories, our deep learning model can help address diagnostic gaps in obstructive HCM and assist in connecting patients to appropriate clinical care.

Limitations of our study include the recruitment and demographics of our study cohort. Both CSMC and SHC are academic tertiary care centers, introducing biases of patient selection. While our algorithm was tested across geographically distinct centers, evaluation in additional care centers across the country should be performed.

Future work can focus on integration of such AI algorithms with ultrasound carts, as additional imaging is needed to interrogate LVOT obstruction. Given the need for real-time feedback for sonographers to initiate further imaging, we anticipate the integration of such an algorithm at the point of scanning for maximal value. By improving disease detection, patients can be connected to disease-specific treatment.

## Data Availability

Model weights will be publicly released on GitHub.

## Acknowledgements

Dr. Ouyang reports support from the National Institutes of Health (R00HL157421, R01HL173487 and R01HL173526) and Alexion, and consulting or honoraria for lectures from EchoIQ, Ultromics, Pfizer, InVision, the Korean Society of Echocardiography, and the Japanese Society of Echocardiography. Ms. Yuan gratefully acknowledges support from the Sarnoff Cardiovascular Research Foundation.

## Notes

### Competing Interest Statement

The authors have declared no competing interest.

### Author Declarations

This study was approved by the Institutional Review Boards at Cedars-Sinai Medical Center and Stanford Healthcare.

### Summary of Updates

This version of the manuscript has been revised to update the author's funding sources.

