## Supplemental Information for "Detection of Left Ventricular Outflow Obstruction from Standard B-Mode Echocardiogram Videos using Deep Learning"

**Supplementary Figure 1.** Distribution of diastolic interventricular septum thickness (IVSd) in a) CSMC case versus control patients and c) SHC case versus control patients **b)** Distribution of reported LVOT gradient in CSMC case patients **d)** Distribution of reported maximum LV velocity in SHC case patients

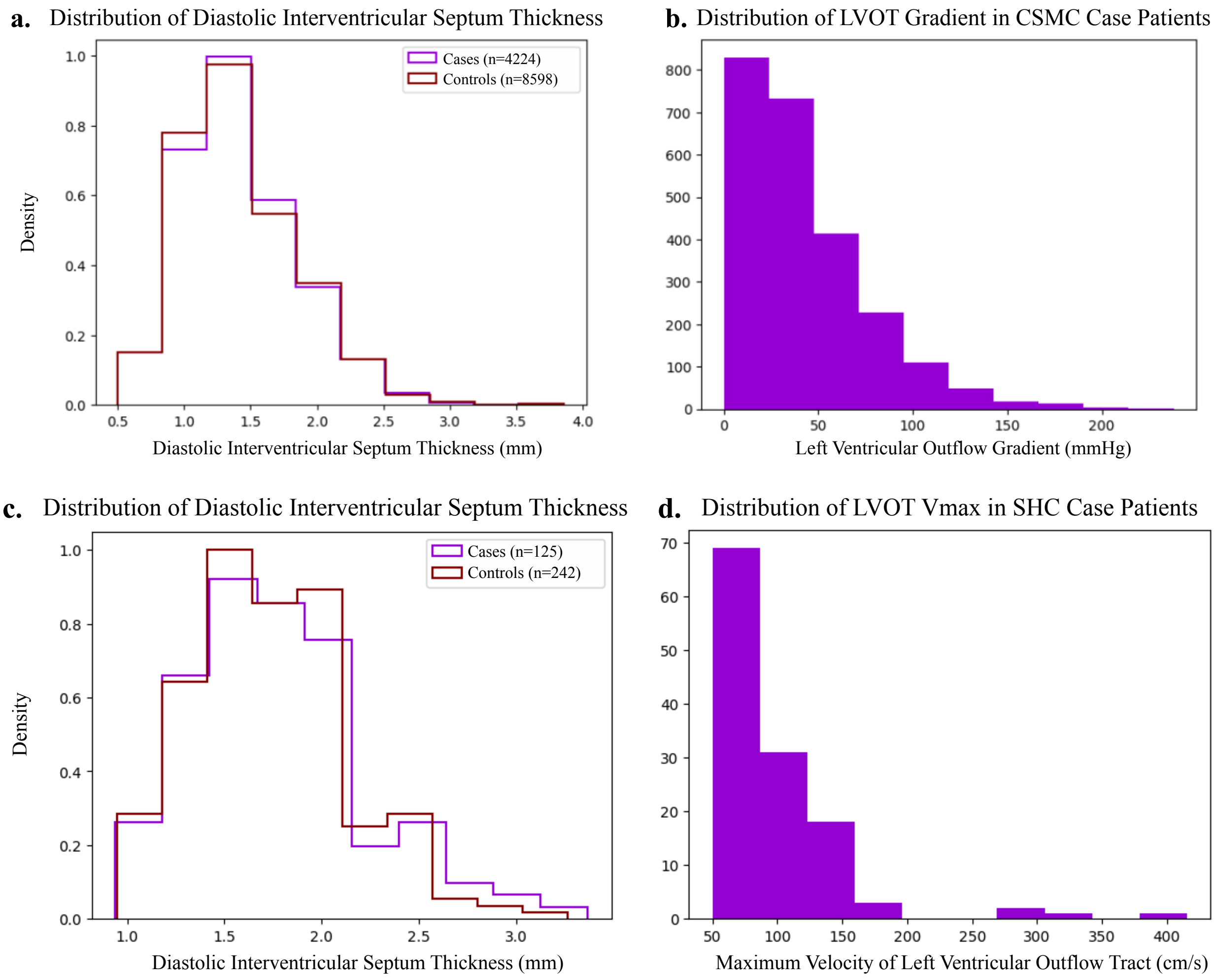

**Supplementary Figure 2**. AUC for subgroups in the CSMC (purple) and SHC (pink) cohorts

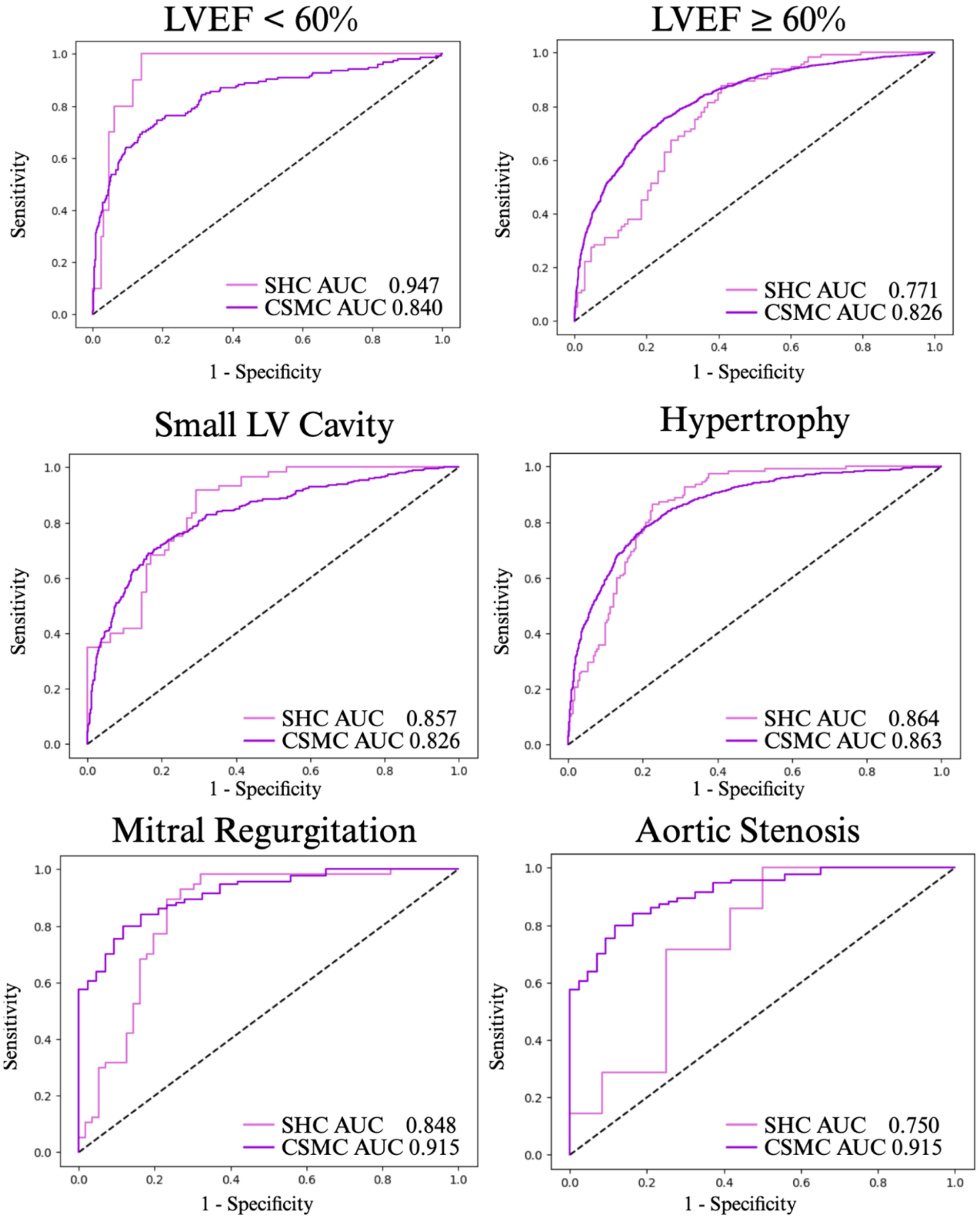

**Supplementary Table 1.** Model performance stratified by patient subgroup

| Subgroup | CSMC Cases | CSMC AUC | SHC Cases | SHC AUC |
|---|---|---|---|---|
| EF <60% | 153 | 0.840 | 10 | 0.947 |
| EF ≥60% | 2023 | 0.826 | 113 | 0.771 |
| Ventricular hypertrophy | 1026 | 0.863 | 125 | 0.864 |
| Small LV cavity | 535 | 0.826 | 60 | 0.857 |
| Aortic stenosis | 94 | 0.915 | 7 | 0.750 |
| Mitral regurgitation | 68 | 0.917 | 57 | 0.848 |

**Supplementary Table 2.** Characteristics of CSMC videos reported as having LVOT obstruction and predicted by the model as having no obstruction (false negatives)

| **False Negative** | |
|---|---|
| **Subgroup** | **Number of Videos** |
| Induced gradient only | 80 (2.6%) |
| SAM without obstruction at rest | 30 (1.0 %) |
| Intracavitary gradient | 103 (3.4%) |
| Ablation study | 6 (0.2%) |
| Left ventricular obliteration | 12 (0.4%) |
| Other | 253 (8.4%) |
| **Total** | **484 (16.0%)** |
| **False Positive** | |
| **Subgroup** | **Number of Videos** |
| Hyperdynamic LV | 142 (8.1%) |
| Hypertrophy | 360 (20.6%) |
| Small LV cavity | 45 (2.6%) |
| Cardiomyopathy | 25 (1.4%) |
| Aortic stenosis | 41 (2.3%) |
| Other | 251 (14.4%) |
| **Total** | **764 (43.7%)** |
